# Integrating multi-tissue expression and splicing data to prioritise anatomical subsite- and sex-specific colorectal cancer susceptibility genes with therapeutic potential

**DOI:** 10.1101/2024.09.10.24313450

**Authors:** Emma Hazelwood, Daffodil M. Canson, Xuemin Wang, Pik Fang Kho, Danny Legge, Andrei-Emil Constantinescu, Matthew A. Lee, D. Timothy Bishop, Andrew T. Chan, Stephen B. Gruber, Jochen Hampe, Loic Le Marchand, Michael O. Woods, Rish K. Pai, Stephanie L. Schmit, Jane C Figueiredo, Wei Zheng, Jeroen R. Huyghe, Neil Murphy, Marc J. Gunter, Tom G. Richardson, Vicki L. J. Whitehall, Emma E. Vincent, Dylan M. Glubb, Tracy A. O’Mara

## Abstract

Numerous potential susceptibility genes have been identified for colorectal cancer (CRC). However, it remains unclear which genes have a causal role in CRC risk, whether these genes are associated with specific types of CRC, and if they have potential for therapeutic targeting. We performed a multi-tissue transcriptome-wide association study (TWAS) across six relevant normal tissues (n=187-670) and applied a causal framework (involving Mendelian randomisation and genetic colocalisation) to prioritise causal associations between gene expression or splicing events and CRC risk (52,775 cases; 45,940 controls), incorporating sex- and anatomical subsite-specific analyses. We identified 35 genes with robust evidence for a potential causal role in CRC, including ten genes not previously identified by TWAS. Among these genes, *SEMA4D* emerged as a significant discovery; it is not located at any established CRC genome-wide association study (GWAS) risk locus and its encoded protein is targeted by an antibody currently being clinically studied for CRC treatment. Several genes showed increased expression associated with CRC risk and evidence of CRC cell dependency in CRISPR screen analyses, highlighting their potential as targets for therapeutic inhibition. A female-specific association with CRC risk was observed for *CCM2* expression, which is involved in progesterone signalling pathways. Subsite-specific associations were also found, including a link between rectal cancer risk and expression of *LAMC1*, which encodes a target for a clinically approved drug. Additionally, we performed a focused analysis of established drug targets to further identify potential therapies for CRC, revealing *PDCD1*, the product of which (PD-1) is targeted by a clinically approved CRC immunotherapy. In summary, our comprehensive analysis provides valuable insights into the molecular underpinnings of CRC and supports promising avenues for therapeutic intervention.

## Introduction

Colorectal cancer (CRC) is the third most common cancer worldwide and the fourth most common cause of cancer-related death^1,2^. There are several established risk factors for CRC, including obesity, alcohol consumption and tobacco use^3–10^ and there is evidence of heterogeneity by sex and anatomical site^10,11^. However, the biological pathways that causally affect CRC development remain poorly understood, which has limited the ability to design suitable therapeutic interventions for prevention and treatment^10,12,13^. Indeed, understanding the genetics underlying disease susceptibility has become an important area of research; drugs with genetic support have been shown to be twice as likely to be successful in clinical trials^14–17^.

Genome-wide association studies (GWAS) have identified common genetic risk variants at over 200 genetic loci associated with CRC risk, including those associated with anatomical subsite-specific CRC^11,18,19^. However, the mechanisms by which these genetic variants affect disease development are generally unknown, hindering translation of these results into clinical applications. Most CRC genetic variants are located outside of coding sequences and their effects are assumed to be mediated through regulation of gene expression, adding complexity to the process of linking variants to their target genes. Given the potential to identify causal disease targets, establishing CRC susceptibility genes from GWAS presents an important opportunity for the development of therapeutic interventions.

Transcriptome-wide association studies (TWAS) are a form of post-GWAS analysis that establishes associations between gene expression and traits. In brief, gene expression is imputed to GWAS of traits of interest (here, CRC risk) using genetic variants which have been previously associated with gene expression in relevant tissues. Given the difficulty in accessing solid tissues for gene expression analyses, TWAS using these tissues are often limited by small sample sizes. S-MultiXcan and joint tissue imputation (JTI) are two TWAS methods which address this issue by incorporating information across multiple tissues to maximise statistical power^20,21^. Including multiple tissues in a single analysis also allows for the identification of the relevant biological tissue for the gene identified – which is important information for drug development. Notably, the S-MultiXcan approach also facilitates analysis of trait associations with alternative splicing events (i.e. processes producing distinct transcripts from the same gene). Alternative splicing is an often neglected mechanism in linking genes to traits despite evidence suggesting that up to ∼30% of GWAS signals may mediate their effects through splicing^22^.

TWAS have successfully identified potential susceptibility genes for many cancers, including breast^23^, endometrial^24^, and CRC^19,25–28^. However, no CRC TWAS performed thus far has stratified by anatomical subsite or sex, which are important aspects of CRC development^9–11,29–31^. Additionally, TWAS for CRC have often lacked a causal framework analysis to account for residual linkage disequilibrium between genetic variants^19,26^. Consequently, it is likely that some previously identified genes represent spurious associations. Identifying genes that causally affect disease development is essential for revealing novel and effective avenues for CRC therapy and treatment.

To address the identified knowledge gaps in CRC, we performed comprehensive multi-tissue expression and splicing TWAS analyses, as outlined in **Figure 1**, with a focus on sex- and anatomical subsite-specific associations. We employed a causal framework performing Mendelian randomization (MR) and genetic colocalisation to identify likely causal associations with CRC risk. Additionally, we evaluated the impact of established drug targets on CRC risk by applying the same framework to 1,163 genes encoding proteins targeted by approved or clinically studied drugs^32^. To prioritise likely causal CRC susceptibility genes for intervention we consulted cancer cell dependency and drug targeting databases. Finally, we evaluated the relationship between four established CRC risk factors (body mass index (BMI), waist:hip ratio (WHR), alcohol consumption, and tobacco use) and identified CRC susceptibility genes.

**Figure 1.**
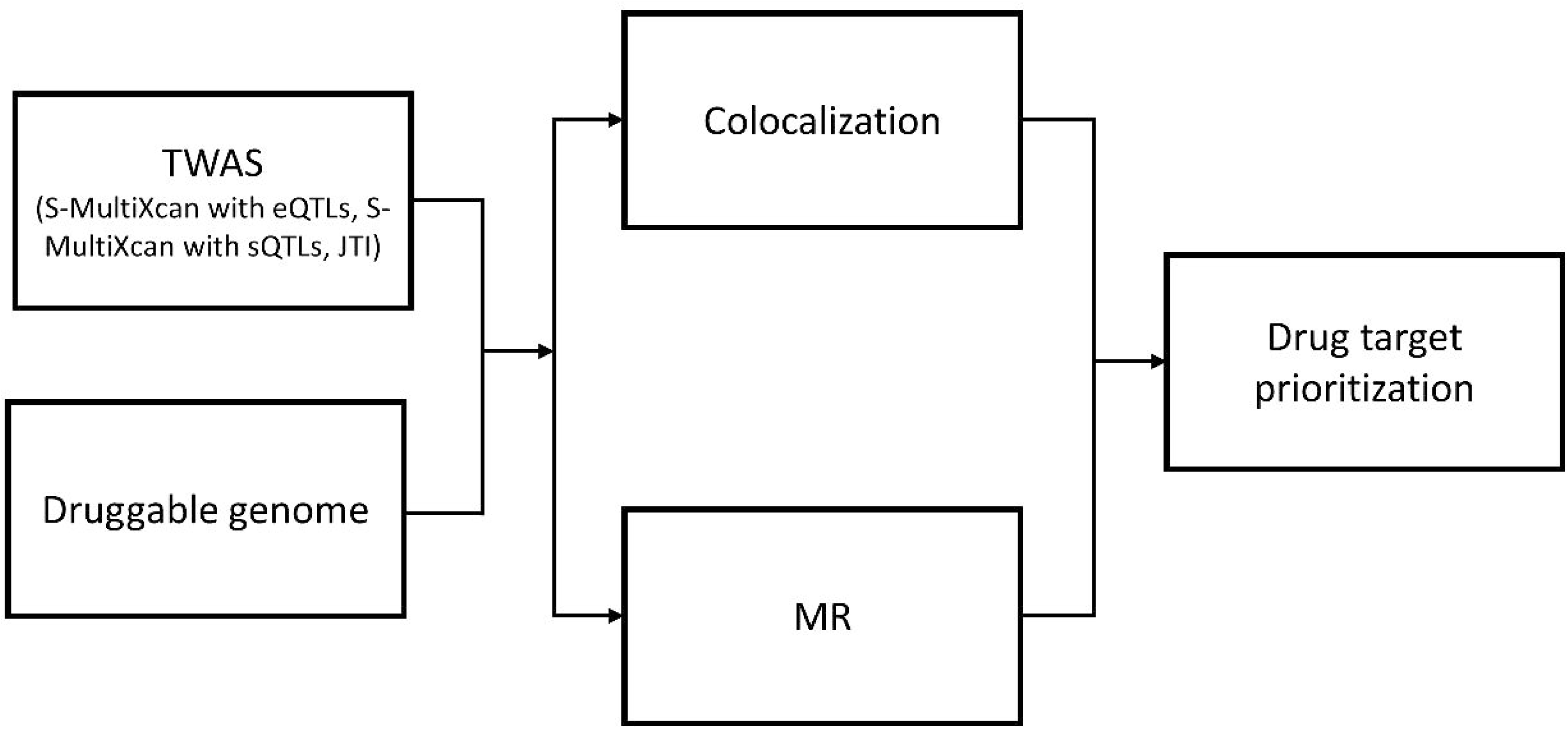
Flowchart detailing analysis plan.

## Results

### Multi-tissue TWAS analyses

To identify genes associated with CRC risk at both the expression and splicing level, we used two multi-tissue TWAS methods: S-MultiXcan and JTI. For S-MultiXcan, we imputed gene expression using expression quantitative trait loci (eQTLs) and splicing events using splicing quantitative trait loci (sQTLs). For JTI we imputed gene expression only as predictive models are not currently available for splicing events. For all TWAS approaches, gene expression or splicing events were imputed using data from the GTEx Project (version 8)^33^. We performed TWAS analyses using data from six tissues previously linked to CRC (subcutaneous and visceral adipose, lymphocytes, and whole blood) or directly relevant to CRC (sigmoid and transverse colon). Associations were tested with risk of overall CRC, as well as sex- or subsite-specific disease. CRC anatomical subsites were defined as per Huyghe et al 2021^11^ (see **Methods**). Briefly, proximal, distal and rectal are mutually exclusive anatomical subsites designated by location of tumour, whereas colon is comprised of proximal colon and distal colon tumours, as well as colon cancer with unspecified location.

Across all three multi-tissue TWAS analyses, 112 unique genes were associated with CRC risk after Bonferroni correction (**Figure 2; Supplementary Tables 1-3**). Of these genes, 68 were identified in the eQTL TWAS analyses, with 30 identified by both JTI and S-MultiXcan approaches. The splicing S-MultiXcan analysis revealed 144 unique splicing events associated with CRC risk, mapping to 60 genes, 23 of which were also identified in at least one of the eQTL TWAS analyses. None of the genes encoding proteins targeted by clinically studied drugs (i.e. ‘druggable genes’) passed correction for multiple testing in any of the TWAS analyses but 772 demonstrated nominal associations (P<0.05).

**Figure 2.**
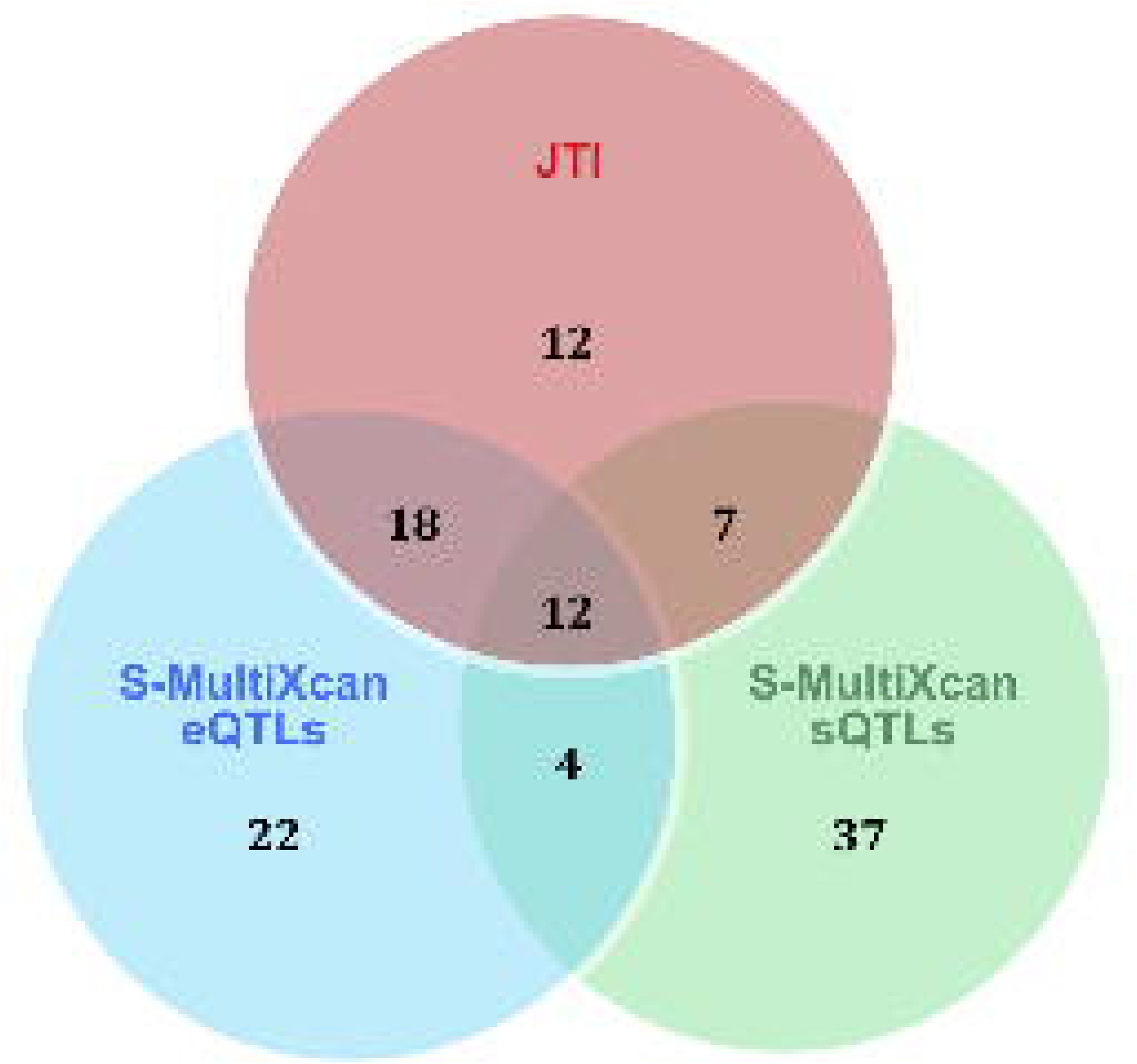
Venn diagram showing overlap of genes identified by each TWAS analysis. JTI = joint tissue imputation; eQTLs = expression quantitative trait loci; sQTLs = splice quantitative trait loci.

### MR analyses

To evaluate the causal effect of gene expression on CRC risk, we performed MR, which uses germline genetic variants as instrumental variables to provide causal estimates (subject to certain assumptions, see **Methods**)^34,35^. Of the 112 genes identified by TWAS, 46 had available cis-genetic variants in at least one of the a priori selected tissues. Among the genes with suitable genetic instruments, 29 passed multiple testing in MR analyses (**Supplementary table 4, Figure 3**). We also included the druggable genes in our causal framework analyses that were nominally associated with CRC risk from TWAS analysis. The expression of seven of these genes passed multiple testing in MR analyses (**Supplementary table 5; Figure 4**).

**Figure 3.**
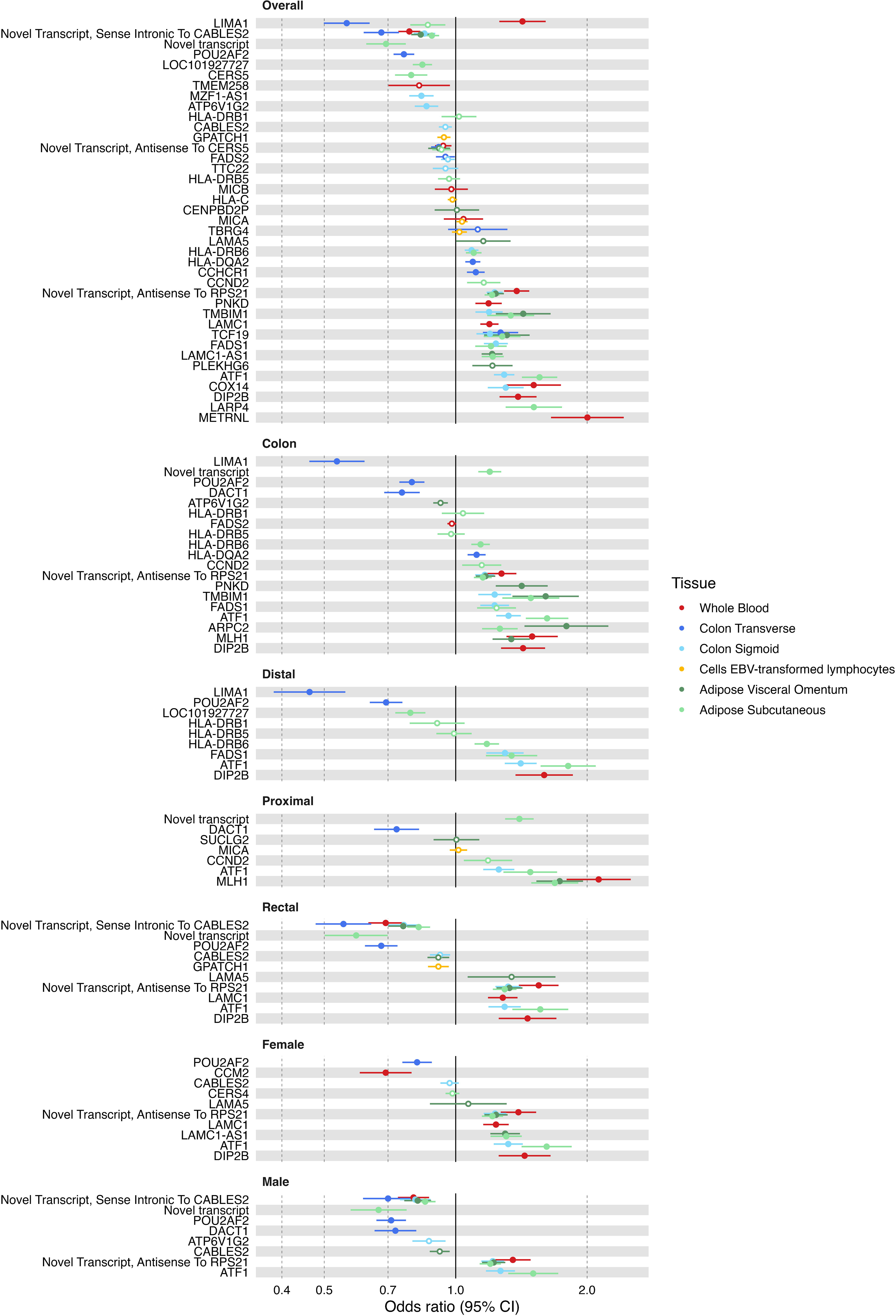
Forest plot showing results from MR analyses of expression of genes from seven tissues identified by TWAS analysis on colorectal cancer risk. Solid points indicate the Bonferroni p-value threshold of P < 4.38 × 10^-5^ (0.05/N*G where N is the number of gene-tissue pairs (161), and G is the number of CRC GWAS (7)) was met in the MR analysis.

**Figure 4.**
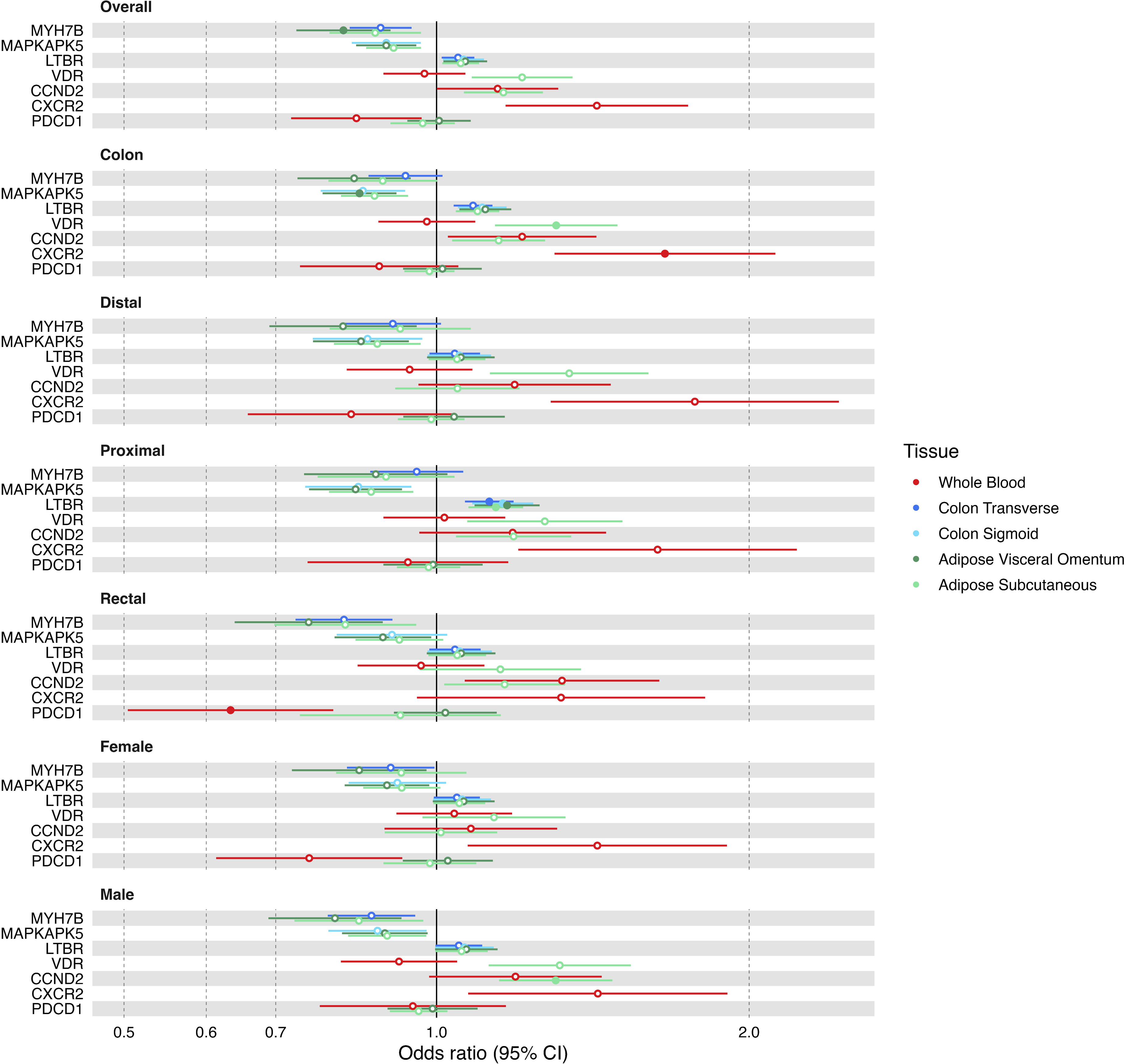
Forest plot showing results from MR analyses of the “druggable genome” on colorectal cancer risk. Only genes which met the Bonferroni P-value threshold of P < 1.32 × 10^-4^ (0.05/number of druggable genes with suitable genetic instruments available (380)) in any MR analysis are shown. Solid points indicate this Bonferroni p-value threshold was met in the MR analysis.

### Colocalisation analyses

Genetic colocalisation analysis can help assess the evidence for causal associations between traits by evaluating whether the same or distinct variant(s) underly the association between two traits^36^. Of the 112 genes identified by TWAS, there was evidence for a shared causal variant between gene expression for 29 of these genes and CRC risk (H_4_, posterior probability of a shared causal variant between the traits, > 0.80; **Supplementary table 6**), and for 19 splicing events that mapped to 12 genes (H_4_ > 0.80; **Supplementary table 7**). Six druggable genes had evidence for a shared causal variant in colocalisation analyses (H_4_ > 0.80) (**Supplementary table 8**).

### Likely causal associations with colorectal cancer risk

To identify likely causal gene associations with CRC risk, we used a stringent framework to prioritise genes: i) passing Bonferroni correction in at least one TWAS analysis; ii) H_4_ > 0.80 in genetic colocalisation analysis; and iii) passing Bonferroni correction in MR analysis or having no suitable genetic instruments available. Using this framework, we identified 35 genes with a likely causal association (Figures 5-7, Table 1). Among these genes, *SEMA4D* is a novel colorectal cancer susceptibility gene, neither located at known colorectal cancer GWAS risk loci nor previously identified by colorectal cancer TWAS. A further nine genes were located at known colorectal cancer GWAS risk loci which had not been previously identified by colorectal cancer TWAS.

**Figure 5.**
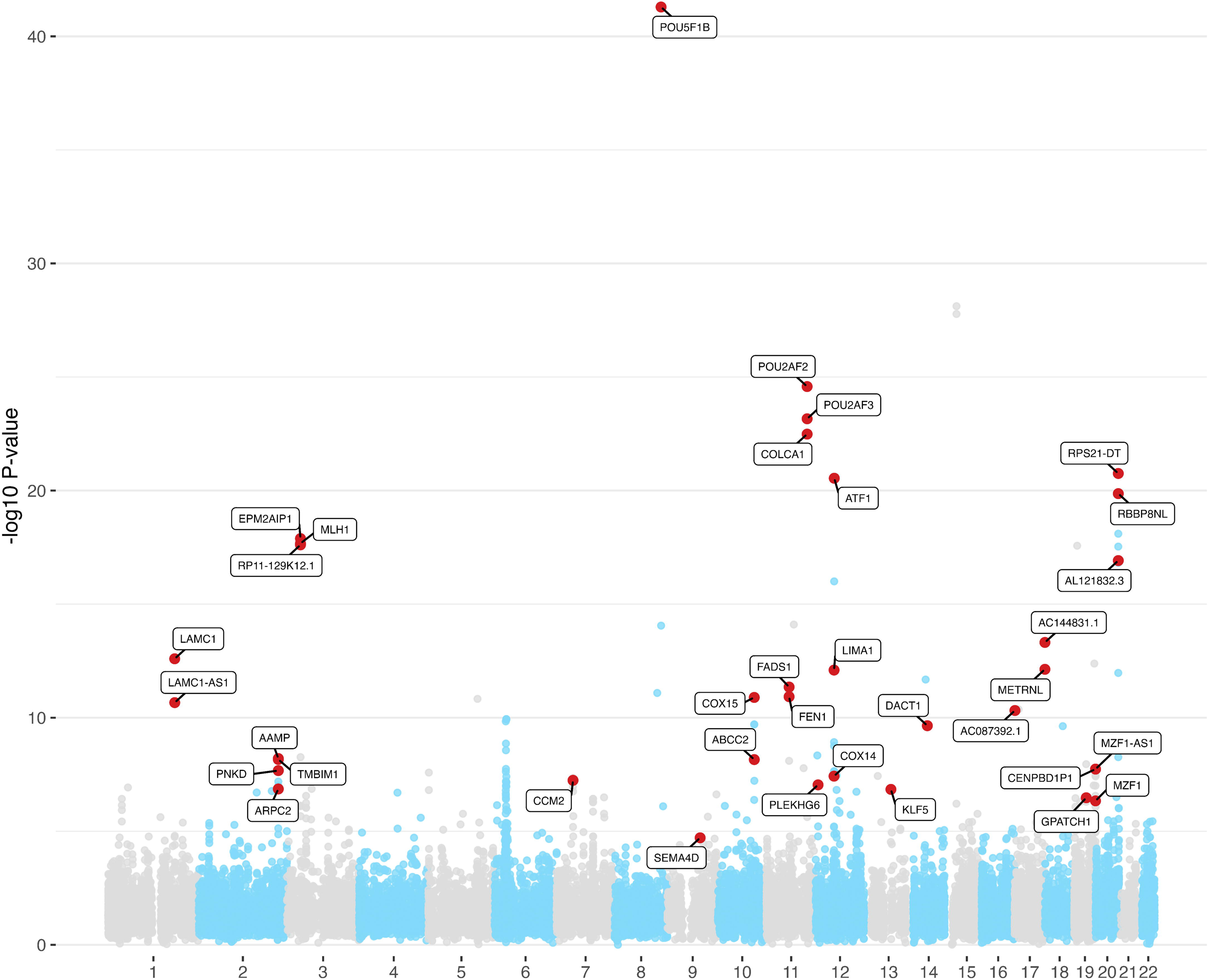
Manhattan plot showing results of S-MultiXcan and JTI TWAS analyses of colorectal cancer risk, for all anatomical subsites combined. Where genes were identified in multiple TWAS analyses, the one with the lowest P-value was retained. Genes labelled are those prioritised following subsequent analyses. Red = prioritised genes for this anatomical subsite; black = prioritised genes for the other anatomical subsite in the Miami plot.

**Figure 6.**
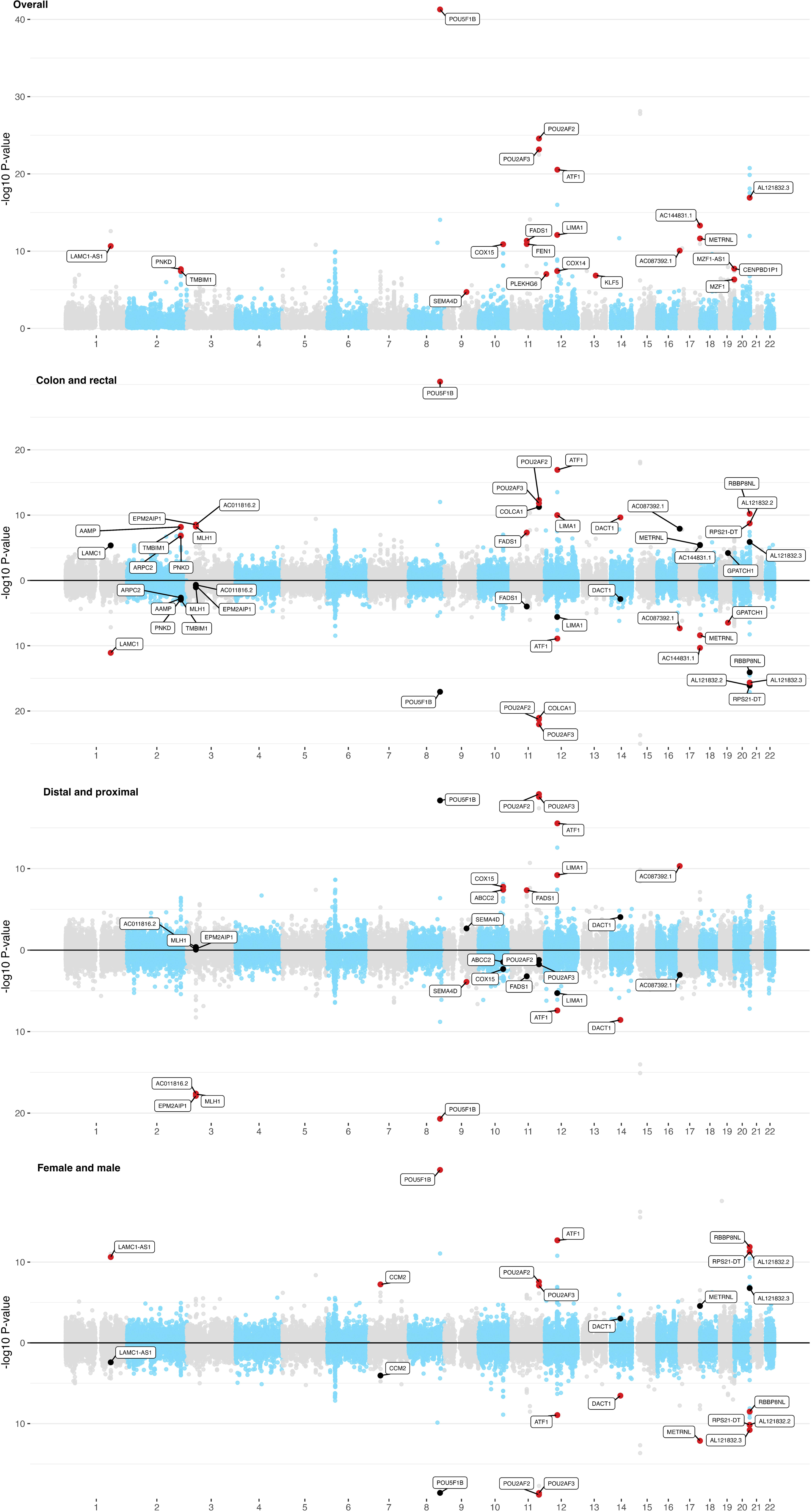
Manhattan and Miami plots showing results of S-MultiXcan and JTI TWAS analyses for each anatomical subsite of colorectal cancer. Where genes were identified in multiple TWAS analyses, the one with the lowest P-value was retained. Genes labelled are those prioritised following subsequent analyses. Red = prioritised genes for this anatomical subsite; black = prioritised genes for the other anatomical subsite in the Miami plot.

**Figure 7.**
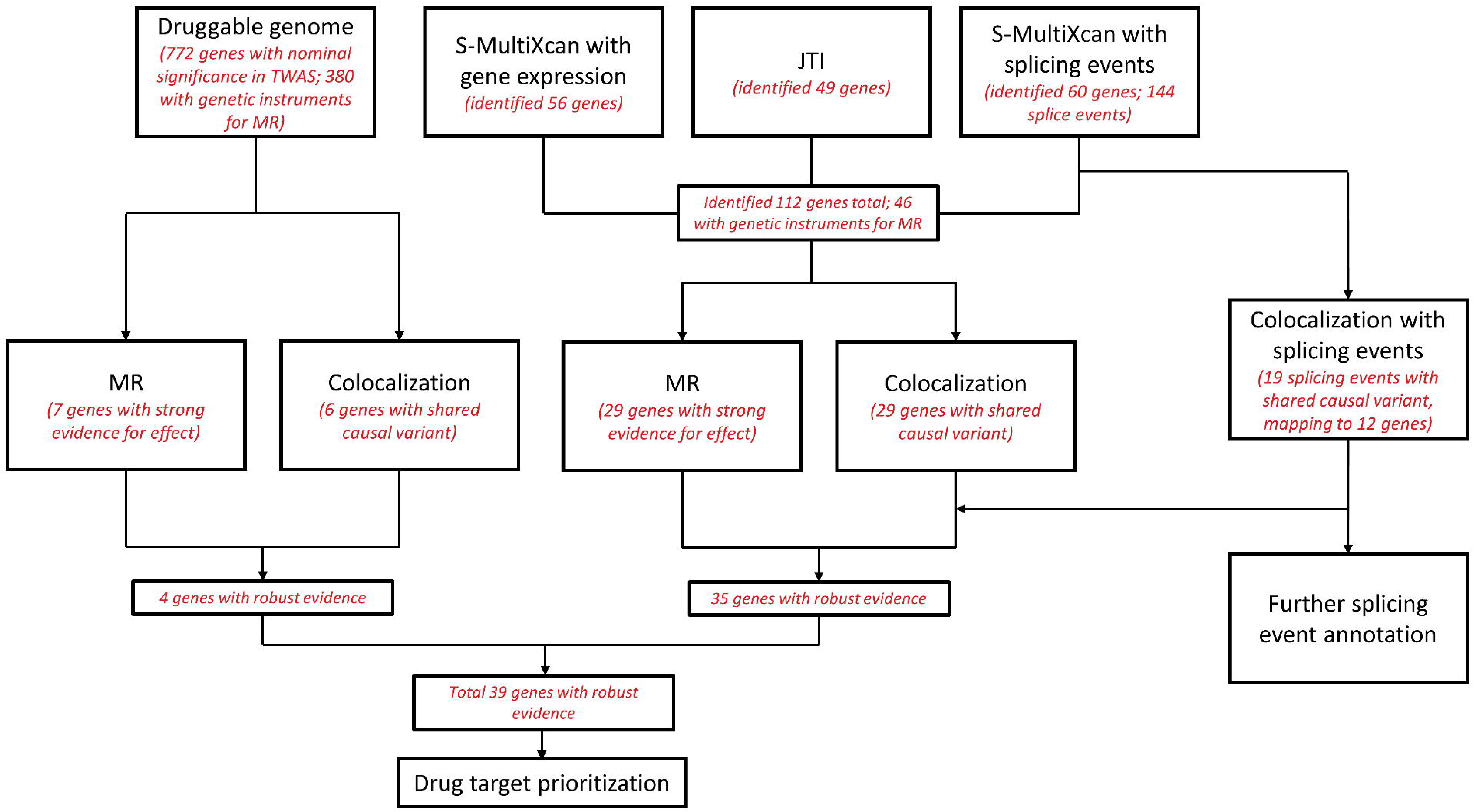
Flowchart showing analysis overview and number of genes/splicing events identified at each stage. “Genes with robust evidence” includes those that had H_4_ above 0.8 in colocalisation analyses, and which either passed Bonferroni correction in the relevant MR analysis (P < 4.38 × 10^-5^; 0.05/N*G where N is the number of gene-tissue pairs (161) and G is the number of CRC GWAS (7) for genes identified in TWAS analyses or P < 1.32 × 10^-4^; 0.05/number of druggable genes with suitable genetic instruments available (380) for genes identified as part of the druggable genome) or which did not have suitable instruments available to be included in the MR analysis. MR = Mendelian randomization.

**Table 1.** Summary table of prioritised genes. Direction = whether the gene was associated with an increased or decreased risk of CRC; Tissue(s) = the tissue(s) in which the gene was associated with CRC risk; Subtype(s) = the CRC subtypes which the gene was associated with; Analysis = the analysis (or analyses) in which the gene was identified, though note that for genes identified in the Splicing MultiXcan analysis the MR analysis, where carried out, evaluated evidence for a causal role of expression (not alternative splicing) of the gene in colorectal cancer risk; Previous GWAS = any previously published CRC GWAS where this locus has been identified; Previous TWAS = any previously published CRC GWAS where this locus has been identified. Tissue abbreviations: AS=adipose subcutaneous; AV=adipose visceral; CS=colon sigmoid; CT=colon traverse; L=EBV-transformed lymphocytes; WB=whole blood. Subtype abbreviations: Overall=all CRC cases; C=colon; R=rectal; P=proximal; D=distal; M=male; F=female. Analysis abbreviations: eJTI=expression JTI; eMX=expression MultiXscan; sMX=splicing MultiXcan.

| Loci | Gene | Ensembl ID | Direction | Tissue(s) | Subtype(s) | Analysis | Previous GWAS (ref)? | Previous TWAS (ref)? |
| --- | --- | --- | --- | --- | --- | --- | --- | --- |
| 2q35 | AAMP | ENSG00000127837 | Inc | AS, AV, CS | C | eJTI | 18 | 19 |
| 10q24.2 | ABCC2 | ENSG00000023839 | Inc | WB | D | eMX | 18 | 19 |
| 17p13.3 | AC087392.1 | ENSG00000262003 | Dec | AS, AV, CT | Overall, D, R | eJTI | 18, 19 | No |
| 17q25.3 | AC144831.1 | ENSG00000261888 | Dec | AS | Overall, R | eJTI, eMX | 18 | No |
| 20q13.33 | AL121832.3 | ENSG00000275437 | Dec | CT | Overall, R, M | eJTI | 18 | 25, 28 |
| 2q35 | ARPC2 | ENSG00000163466 | Inc | AS, AV | C | sMX (MR expression) | 18 | 19, 28 |
| 12q13.12 | ATF1 | ENSG00000123268 | Inc | AS, AV, CS, CT | All analyses | eJTI, eMX, sMX | 18 | 19, 28 |
| 7p13 | CCM2 | ENSG00000136280 | Dec | WB | F | eJTI | 18 | 19 |
| 19q13.43 | CENPBD2P | ENSG00000213753 | Unk | AS | Overall | sMX | 18 | No |
| 11q23.1 | COLCA1 | ENSG00000196167 | Dec | CT | R | eJTI | 18 | 19, 25, 28 |
| 12q13.12 | COX14 | ENSG00000178449 | Inc | AS | Overall | eJTI | 18 | 19, 28 |
| 10q24.2 | COX15 | ENSG00000014919 | Unk | CT, L | Overall, D | sMX | 18 | 19 |
| 14q23.1 | DACT1 | ENSG00000165617 | Dec | CT | C, P, M | eJTI, eMX | 18 | 19, 25, 28 |
| 3p22.2 | EPM2AIP1 | ENSG00000178567 | Inc | AV, WB | C, P | eJTI, sMX | 11 | No |
| 11q12.2 | FADS1 | ENSG00000149485 | Inc | AS, CS | Overall, C, D | eJTI, eMX, sMX | 18 | 19 |
| 11q12.2 | FEN1 | ENSG00000168496 | Inc | AS | Overall | eMX | 18 | 19 |
| 19q13.11 | GPATCH1 | ENSG00000076650 | Unk | L | R | sMX | 18 | 19, 28 |
| 2q35 | GPBAR1 | ENSG00000179921 | Inc | WB | Overall | Druggable genome | 18 | 19 |
| 13q22.1 | KLF5 | ENSG00000102554 | Unk | CT | Overall | sMX | 46 | 19 |
| 1q25.3 | LAMC1 | ENSG00000135862 | Inc | WB | R | eJTI | 18 | 19, 28 |
| 1q25.3 | LAMC1-AS1 | ENSG00000224468 | Inc | L | Overall, R | eJTI | 18 | No |
| 12q13.12 | LIMA1 | ENSG00000050405 | Dec | CT | Overall, C, D | eJTI, eMX | 18 | 19 |
| 12p13.31 | LTBR | ENSG00000111321 | Inc | AV, CS, CT | P | Druggable genome | 18 | 28 |
| 17q25.3 | METRNL | ENSG00000176845 | Inc | AV, WB | Overall, R, M | eJTI, sMX | 18 | 28 |
| 3p22.2 | MLH1 | ENSG00000076242 | Inc | AS, AV, L, WB | C, P | eJTI, eMX, sMX | 11 | No |
| 19q13.43 | MZF1 | ENSG00000099326 | Unk | CS | Overall | sMX | 18 | 19, 28 |
| 19q13.43 | MZF1-AS1 | ENSG00000267858 | Dec | CS | Overall | sMX (MR expression) | 18 | No |
| 2q37.3 | PDCD1 | ENSG00000188389 | Inc | WB | R | Druggable genome | No | No |
| 12p13.31 | PLEKHG6 | ENSG00000008323 | Unk | CT | Overall | sMX | 18 | 19 |
| 2q35 | PNKD | ENSG00000127838 | Inc | AV, WB | Overall, C | eJTI, eMX | 18 | 19, 28 |
| 11q23.1 | POU2AF2 | ENSG00000150750 | Dec | CT | Overall, C, R, D, M, F | eJTI, eMX | 18, 109 | 19, 25, 28, 109 |
| 11q23.1 | POU2AF3 | ENSG00000214290 | Dec | CS, CT | Overall, C, R, D, M, F | eJTI, eMX, sMX | 18, 109 | 19, 25, 28 |
| 8q24.21 | POU5F1B | ENSG00000212993 | Dec | CT | Overall, C, P, F | eJTI, eMX | 18, 109 | 19, 25, 109 |
| 1p31.1 | PTGER3 | ENSG00000050628 | Inc | AV | C, P | Druggable genome | 11 | No |
| 20q13.33 | RBBP8NL | ENSG00000130701 | Dec | CT | C, F, M | eJTI, eMX | 18 | 25, 28 |
| 3p22.2 | RP11-129K12.1 | ENSG00000272334 | Inc | AS, CT | C, P | eJTI, eMX | 11 | No |
| 20q13.33 | RPS21-DT | ENSG00000273619 | Inc | All tissues | C, F, M | eJTI | 18 | 19 |
| 9q22.2 | SEMA4D | ENSG00000187764 | Unk | L | Overall, P | sMX | No | No |
| 2q35 | TMBIM1 | ENSG00000135926 | Inc | AS, AV, CS | Overall C | eJTI | 18 | 19, 25, 28 |

The analysis revealed several context-specific associations. Twenty likely causal susceptibility genes were identified solely through associations with expression and nine through associations with splicing alone. Furthermore, of the 35 likely causal genes, 21 showed tissue-specific associations (i.e. associations unique to expression or splicing in one tissue): four genes were found through analysis of subcutaneous adipose, two through sigmoid colon, nine through transverse colon, three through lymphocytes and three through whole blood. Regarding anatomical subsites, two genes were exclusively associated with colon cancer risk (*AAMP* and *ARPC2*), three genes with both colon and proximal colon cancer risk (*EPM2AIP1, MLH1 and RP11-129K12.1*), one with distal colon cancer risk (*ABCC2*) and three with rectal cancer (*COLCA1*, *LAMC1* and *GPATCH1*) risk. For all but *AAMP*, differences in TWAS effect sizes for these genes were observed between subtypes (**Supplementary Figures 1 & 2**). Lastly, one gene was specifically associated with female colorectal cancer risk (*CCM2*; Supplementary Figure 1N).

For the analysis of the druggable genes, we conducted an exploratory analysis by focussing on genes that were nominally significant in at least one TWAS analysis. To prioritise genes for causality, we selected those passing H_4_ > 0.80 in genetic colocalisation analysis and Bonferroni-correction in MR analysis. This approach revealed four genes (*GPBAR1, LTBR, PDCD1* and *PTGER3*) (**Figures 4 and 7, Table 1**).

### Splicing event annotation

To provide further support for likely causal splicing associations, we explored underlying splicing mechanisms. Using a bioinformatic splicing pipeline to analyse CRC GWAS risk variants for effects on the likely causal splicing events, we found that a splicing event related to PLEKHG6 (intron_12_6317696_6317899; Supplementary Table 9) could be explained by rs1468603 (chr12:6317886 C>T). Specifically, the T allele was predicted to activate an exonic cryptic acceptor, enhancing the inclusion of an in-frame 45 bp deletion of PLEKHG6 (NM_001384598.1) exon 10, corresponding to the intron_12_6317696_6317899 splicing event.

### Evaluating drug targeting opportunities provided by likely causal susceptibility genes

In addition to specifically analysing druggable targets, we investigated the druggability of proteins encoded by the likely causal susceptibility genes using the Pharos and Open Targets platforms. These databases identified proteins encoded by *LAMC1* and *SEMA4D* as targets of clinically studied drugs. Laminin subunit gamma 1, encoded by *LAMC1*, is degraded by ocriplasmin, a recombinant proteinase drug used to treat vitreomacular adhesion. *SEMA4D* encodes semaphorin 4D which is inhibited by pepinemab, an antibody that has been clinically studied for treatment of several cancer types, including a phase I trial of CRC (Clinicaltrials.gov: NCT03373188). We also identified five genes (*ABCC2, ATF1, FADS1, FEN1* and *KLF5*) whose protein products bind to small molecules, supporting their potential druggability.

We evaluated the potential for efficacy in therapeutic targeting of likely causal susceptibility genes by assessing if their expression is required for CRC cell line viability. Using the BioGRID Open Repository on CRISPR Screens, we found CRC cell lines were dependent on 14 of the likely causal susceptibility genes (Supplementary Table 10). Among these genes, ten were identified through expression TWAS approaches (Table 1). Consistent with the dependency findings, increased expression of seven genes, including *AAMP* and *FEN1*, associated with CRC risk. *AAMP* showed particularly consistent findings, with CRC cell lines demonstrating dependency for *AAMP* expression in 80% of the studies in which it was tested. CRC cell lines also showed frequent dependency for expression of FEN1 (48% of studies), which encodes a potentially druggable protein.

### Shared causal pathways with known CRC risk factors

To investigate whether the likely causal susceptibility genes may relate to known CRC risk factors, we performed genetic colocalisation. We evaluated evidence for a shared causal variant between the expression of 28 prioritised genes (i.e. not including the seven genes that had robust evidence for splicing only) and each of four established CRC risk factors - BMI, WHR, alcohol consumption, and smoking initiation. Among these genes, we found evidence of colocalisation (posterior probability of H_4_ > 0.80) for two genes (*AAMP* and *TMBIM1*) with WHR (**Supplementary Table 11**).

## Discussion

Our analysis combined two multi-tissue TWAS methods with a causal framework to identify CRC susceptibility genes. Through this framework, we prioritised 35 genes with strong evidence for a causal role in colorectal cancer risk, with associations extending to specific disease subtypes and expression in distinct tissues. In addition, our analysis of the druggable genome revealed four genes with suggestive evidence for a causal role in colorectal cancer risk. The subsequent drug target analyses allowed us to highlight candidates for future investigation.

While previous TWAS for CRC have been conducted, these analyses have not been stratified by anatomical subsite or sex, which are important aspects of CRC aetiology. The importance of stratified analysis is demonstrated by our findings for a causal role of *CCM2* in female-specific colorectal cancer. Cerebral cavernous malformation 2 (*CCM2*) is a component of the CCM signalling complex, which has a role in regulating several signalling cascades, including progesterone signalling^37,38^. This complex has previously been implicated in tumourigenesis, in particular in breast cancer through its role in sex hormone regulation, supporting our findings of a potential sex-specific role for *CCM2*^37,38^. In addition our findings support evidence from GWAS that genes at locus 3p22.2 (including MLH1 and EPM2AIP1) have proximal colon cancer-specific effects^11,39–43^. Though loss of function MLH1 variants are known to be associated with proximal colon cancer, we found that increased MLH1 expression was associated with increased cancer risk. A similar, albeit nominally significant TWAS finding was previously reported ^28^. Further research is thus required to understand the direction of effect of MLH1 expression on proximal colon cancer risk.

Among the likely causal genes, *SEMA4D* emerged as a novel CRC susceptibility gene, neither located at known CRC GWAS risk loci nor previously identified by CRC TWAS. *SEMA4D* was identified through association of its alternative splicing with colorectal cancer risk, highlighting the importance of studying this mechanism using TWAS approaches. *SEMA4D* encodes a protein with immunoregulatory activity^44^, consistent with its association with CRC risk through splicing effects in lymphocytes. *SEMA4D* has been assessed as a target for CRC treatment due to its association with progression and angiogenesis^45^ with findings from a phase I trial of an antibody antagonist not yet reported. A further nine genes, located at known CRC GWAS risk loci, had not been previously identified by CRC TWAS. These findings may possibly be due to the lack of anatomical subsite-stratified analyses in previous TWAS (e.g. *EPM2AIP1* and *MLH1*, which both showed evidence for colon-specific effects in our analyses), or our inclusion of alternative splicing events (e.g. *CENPBD2P* and *MZF1-AS1*).

*LAMC1* emerged as another likely causal susceptibility gene encoding a target (laminin gamma subunit 1) of a clinically studied drug (ocriplasmin). *LAMC1* has previously been identified as a CRC susceptibility gene through GWAS and other approaches^19,46–49^. The laminin family of proteins are key components of the basal membrane and have been implicated in CRC progression^50,51^. We found genetically predicted increased expression of *LAMC1* was associated with increased rectal cancer risk, providing support for therapeutic inhibition of *LAMC1*. Ocriplasmin, a synthetic form of plasmin which targets laminin, is currently used to treat eye-related diseases and is also in phase II trials for several other conditions, including stroke and deep vein thrombosis^52–54^. While prior research has suggested ocriplasmin as a candidate drug for CRC treatment^55^, further drug development would be required due to the current need for direct injection of ocriplasmin and its moderate stability^56^.

We found that most likely causal susceptibility genes (e.g. AAMP and FEN1) for which CRC cell lines have shown dependency align with TWAS findings where increased expression was associated with increased CRC risk. This alignment underscores their potential for effective therapeutic intervention. The most consistent findings of CRC dependency were for AAMP which encodes angio-associated migratory cell protein (AAMP), with a role in angiogenesis, cell migration^57^, and CRC metastasis^58^. We also found evidence for colocalisation of AAMP expression with WHR suggesting that AAMP may also impact CRC risk through effects on adipose distribution, or vice versa. Although there are no current inhibitors of AAMP, Open Targets indicates there is potential for inhibition through antibody or protein targeting chimera approaches. FEN1 also demonstrated consistent CRC dependency. The metallonuclease encoded by FEN1 has a role in DNA replication and double-strand break repair^59^. Promisingly, FEN1 small molecule inhibitors have been developed that show anti-cancer effects in experimental models^60^.

We also performed a comprehensive analysis of the “druggable genome”^32^. We focussed on genes that were nominally significant in at least one TWAS analysis and prioritised genes with evidence of genetic colocalisation (H_4_ > 0.80) with CRC risk and which met the Bonferroni-correction in an MR analysis. This revealed suggestive evidence for a causal effect of expression of four genes (*PDCD1, GPBAR1, PTGER3 and LTBR*) on CRC risk. Among these, there were two tissue-specific associations observed in whole blood (*GPBAR1* and *PTGER3*). Additionally, we found associations with unique anatomical subsite cancers: LTBR with risk of proximal colon cancer and *PDCD1* with risk of rectal cancer. *PDCD1* encodes programmed cell death 1 (PDCD-1 or PD-1) protein, which is targeted by inhibitors used to treat microsatellite instability-high or mismatch repair-deficient metastatic CRC^61–66^. Our TWAS and MR analyses suggested that increased (rather than decreased, replicating the use of an inhibitor) expression of *PDCD1* reduced risk of rectal cancer. This conflicts with evidence that PDCD-1 suppresses the immune system’s ability to destroy cancer cells, as one would assume that in this case increased *PDCD1* expression would increase (not decrease) cancer risk^67^. However, we note that we only see strong evidence for a causal role of *PDCD1* expression in blood (not colon tissue) on cancer risk – suggesting that the mechanism linking *PDCD1* expression and colorectal cancer risk may be more complex than the presumed local effects within colorectal tissue. PTGER3 encodes a receptor for prostaglandin E2 that is targeted by misoprostol, an approved drug for gastric ulcers and reflux disease and which has shown efficacy in colon cancer xenograft models^68^. We replicated previous GWAS evidence that PTGER3 may have a role in proximal colon cancer and may be less relevant to rectal cancer^11^. LTBR encodes the tumour necrosis factor receptor lymphotoxin beta receptor (LTBR) which is targeted by an antibody agonist^69^. However, an antibody antagonist is likely to be required for effective treatment given increased LTBR expression in several tissues was associated with risk of proximal colon cancer.

Our analysis aimed to robustly prioritise genes for CRC susceptibility by using multiple tissues alongside a causal framework. However, the sample sizes for available data for TWAS analyses are still relatively small compared to the CRC GWAS, which potentially impacts our ability to genetically predict gene expression and detect associations with CRC risk. In addition, our analyses were limited to genes with expression that can be predicted using available TWAS models, meaning some potentially casual genes may not be captured in our analyses. Additionally, many of our MR analyses were restricted to a single SNP, meaning we were unable to employ various “pleiotropy-robust” models to evaluate exclusion restriction assumptions. The colocalisation analyses performed here maintain the single causal variant assumption, which assumes that there are not multiple causal variants in the given genomic region for the traits. This may not be true in some cases, which may mean that some of the genes which we identified as not having strong evidence for a causal relationship with CRC risk are false negatives. Furthermore, we did not evaluate the sensitivity of our colocalisation analyses to alternative window sizes or prior probabilities, which are important aspects of colocalisation analyses^15,70^. Our study also presents further limitations that could be addressed in future research: i) our analysed were restricted to individuals of predominantly European ancestries, which limits the generalisability of our findings to other populations and contexts; ii) the MR analyses performed here assume linearity between gene expression and CRC risk, which may not capture more complex interactions and non-linear relationships; iii) the use of available summary data limited our ability to perform analyses with sex-specific gene expression data that could provide insights into differential CRC risk.

## Conclusion

Given the increase in CRC worldwide, understanding the biological mechanisms leading to carcinogenesis is becoming increasingly important^71^. Additionally, as more screening programmes are rolled out globally, opportunities to prevent CRC development in high-risk individuals are also increasing. Therefore, the identification of novel pharmaceutical targets for the prevention and treatment of this disease remains a priority. Our analyses have identified genes with robust evidence for a potential causal role in CRC development, providing insights into its aetiology and presenting tangible opportunities for targeted therapeutic interventions.

## Methods

### CRC GWAS

Supplementary Table 12 shows the GWAS used in all analyses. Summary genetic association data for CRC risk (52,775 cases, 45,940 controls) were obtained from a meta-analysis of the Colorectal Transdisciplinary Study (CORECT), the Colon Cancer Family Registry (CCFR), and the Genetics and Epidemiology of CRC (GECCO) consortium^11,18^. Summary genetic association data were obtained stratified by site (colon, 28,736 cases; proximal colon, 14,416 cases; distal colon, 12,879 cases; and rectal, 14,150 cases; 43,099 controls) and sex (female, 24,594 cases, 23,936 controls; male, 28,271 cases, 22,351 controls). Colon cancer included proximal colon (any primary tumour arising in the cecum, ascending colon, hepatic flexure, or transverse colon), distal colon (any primary tumour arising in the splenic flexure, descending colon or sigmoid colon), and colon cases with unspecified site. Rectal cancer included any primary tumour arising in the rectum or rectosigmoid junction. CRC was classified using ICD-10 codes and most cases were incident CRC. All participants in the anatomical subsite-specific CRC analyses were of European ancestries, and approximately 92% of participants in the overall CRC GWAS were European (∼8% were East Asian). Imputation of GWAS summary statistics was performed using the Michigan imputation server and HRC r1.0 reference panel. Regression models were adjusted for age, sex, genotyping platform, and genomic principal components as described previously^18^. Ethics were approved by respective institutional review boards.

### Multi-tissue TWAS analyses

To identify genes with expression or splicing events associated with CRC risk, we utilised two multi-tissue TWAS methods. First, we performed S-MultiXcan^20^, which is an extension of S-PrediXcan^72^. Briefly, S-PrediXcan identifies genes with expression or splicing events that are associated with a phenotype of interest using linear prediction models to impute gene expression and splicing events to the trait GWAS. We performed S-PrediXcan using precomputed gene expression or alternative splicing prediction models and linkage disequilibrium (LD) reference datasets, downloaded from the PredictDB data repository (http://predictdb.org/). S-MultiXcan extends this approach by incorporating gene expression prediction across multiple tissues using multivariate regression. Effect sizes were calculated using multivariate adaptive shrinkage^73^, which is a flexible statistical approach that leverages information on the similarity between variables to improve effect estimation. This approach was applied to variants identified by fine-mapping using deterministic approximation of posteriors^74,75^, which performs joint enrichment analysis of GWAS and quantitative trait loci data to annotate genetic variants. Given that these models often rely on variants that may be absent from most trait GWAS, we performed additional harmonization and imputation of the CRC GWAS prior to these analyses, as recommended by the S-MultiXcan authors. We performed the S-PrediXcan and S-MultiXcan analyses for both eQTLs and sQTLs. For the S-MultiXcan splicing analysis, splice events were mapped to relevant genes using the GTEx splicing mapping file (downloaded from www.gtexportal.org/home/datasets).

Second, we performed JTI as another means to identify genes with expression associated with CRC^21^. This method is another extension of S-PrediXcan and again imputes gene expression to trait GWAS by incorporating information across multiple tissues to improve prediction quality. We performed JTI using precomputed models for gene expression imputation which exploit measures of similarity between tissues based on expression data and cell-specific regulatory elements. The pretrained JTI models were downloaded from Zenodo (https://doi.org/10.5281/zenodo.3842289).

Both TWAS methods incorporate information about gene expression or splicing events across multiple biological tissues to maximise statistical power. As the architecture of eQTLs and sQTLs can differ substantially across tissues^76^, previous evidence has suggested that using only those from tissues which are mechanistically related to the GWAS trait can avoid spurious findings^77^. Thus, for both TWAS methods, we used data (from GTEx Project (version 8)^33^) from six biologically relevant tissues for CRC: two adipose tissue types (subcutaneous adipose (n=581) and visceral (omentum) adipose (n=469)), which may capture important adiposity-related CRC pathways^10^; two colon tissue types (transverse colon (n=368) and sigmoid colon (n=318)), which may capture locally important oncogenic processes; one immune tissue type (Epstein-Barr virus-transformed lymphocytes (n=187)), given recent links between circulating white blood cells and CRC risk^78^; and whole blood (n=670), which may capture a range of clinically important circulating factors. We removed variants with a minor allele frequency (MAF) < 1% from the CRC GWAS summary statistics prior to TWAS analyses.

Given our aim of identifying genes which should be prioritised in future CRC research, for all TWAS analyses we applied a Bonferroni-correction to identify genes associated with CRC risk (0.05/(N*G*T), where N is the number of genes or splice events included in the analysis, G is the number of CRC GWAS tested (overall, female, male, colon, distal, proximal, rectal), and T is specific to the JTI analyses and is the number of tissues included in the analysis (of subcutaneous adipose tissue, visceral adipose tissue, transverse colon, sigmoid colon, lymphocytes, and whole blood). Any genes passing this Bonferroni threshold in at least one of the analyses (P < 3.91 × 10^-7^ in S-MultiXcan eQTL analysis; P < 5.49 × 10^-7^ in S-MultiXcan sQTL analysis; P < 6.01 × 10^-8^ in JTI analysis) were taken forward to the MR analyses.

Full S-PrediXcan results are available for download from Zenodo (https://doi.org/10.5281/zenodo.12805739).

### MR analyses

MR is a genetic epidemiological approach which, under certain assumptions, can estimate causal effects between phenotypes in observational settings^34,35^. MR uses germline genetic variants as instrumental variables for exposures. Since these variants are randomly assorted at meiosis and fixed at conception, MR analyses should be less prone to confounding by environmental factors and reverse causation bias than conventional observational studies. The three core assumptions of MR state that: (i) the genetic variant(s) are strongly and robustly associated with the exposure; (ii) there is no confounding of the genetic variant(s)-outcome relationship (e.g., population stratification); (iii) the genetic variant(s) only affect the outcome through their effect on the exposure.

We performed MR to evaluate evidence for a causal effect of tissue-specific gene expression for all genes identified in the TWAS analyses on the relevant CRC outcome (46 out of 112 genes were instrumentable). Summary genetic data for gene expression (i.e. eQTLs) were obtained from GTEx (version 8)^33^. We identified genetic instruments as genetic variants which are cis-acting (i.e. within 100kb of the gene coding region), strongly associated with gene expression (P < 5 × 10^-8^), independent (r^2^ < 0.001), and had an F-statistic > 10. Where only a single genetic variant was available, we calculated the Wald ratio to generate effect estimates; where multiple genetic variants were available, an inverse variable weighted (IVW) multiplicative random effects model was used. A Bonferroni-correction was applied to account for multiple testing (P < 4.38 × 10^-5^; 0.05/N*G where N is the number of gene-tissue pairs (161) and G is the number of CRC GWAS (overall, male, female, colon, distal, proximal, rectal)).

In addition to genes identified through the TWAS analysis, given our focus on identifying genes which hold high therapeutic potential for CRC prevention, as an additional analysis we also performed MR to evaluate evidence for a causal role of expression of 380 (out of 1,163) previously identified actionable drug targets in CRC development^32,79^ that had nominal significance in at least one TWAS analysis. We used the same genetic instrument identification process as with the prior MR analysis and applied a Bonferroni correction to account for multiple testing (P < 1.32 × 10^-4^; 0.05/number of druggable genes with suitable genetic instruments available (380)).

All genetic variants used in MR analyses are available in Supplementary Table 13. A completed MR-Strobe^80,81^ checklist is available in the Supplementary note (downloaded from: https://www.strobe-mr.org/).

### Colocalisation analyses

Genetic colocalisation uses a Bayesian framework to determine whether the causal variant(s) within a locus relating to multiple phenotypes is shared between the traits^36^. This shared causal variant is necessary (but not sufficient in the absence of other evidence) for a causal relationship. We performed genetic colocalisation under the single causal variant assumption^82^ of (i) gene expression (eQTL) and CRC for all genes which were identified by any of the TWAS analyses and the relevant CRC anatomical subsite; (ii) gene expression (eQTL) and CRC for all genes from the aforementioned “druggable genome” for which data were available and all CRC anatomical subsites; and (iii) gene splicing (sQTL) and CRC for all genes identified in the S-MultiXcan splicing analysis and the relevant CRC anatomical subsite. Colocalisation was performed using the priors p1 = 1 x 10^-4^, p2 = 1 x 10^-4^, and p12 = 1 x 10^-5^, with all genetic variants within 100kb of the relevant gene coding region^82,83^. A posterior probability of >0.80 for H_4_ was used to indicate strong evidence for a shared causal variant, and thus evidence for a causal relationship, between the traits^84^.

### CRC dependency

To determine the dependency of CRC cell lines on likely causal susceptibility genes, we interrogated the BioGRID Open Repository of CRISPR Screens (https://orcs.thebiogrid.org/) and identified genes whose knockout impacts cell viability, using the study authors’ defined threshold for evidence of gene dependency.

### Open Targets database

We used the Open Targets (https://www.targetvalidation.org) and Pharos (https://pharos.nih.gov/) platforms to evaluate drug target tractability and to identify drugs which may target the products of genes identified in our analysis.

### Splicing event annotation

We employed the SpliceAI-10k calculator to investigate downstream consequences of splice events, which has been described previously^85^. The SpliceAI calculator uses deep neural networks trained on large-scale catalogues of alternative transcript isoforms and tissue sQTLs to assess splicing variants for their likely splicing effects (i.e., 511 and 311 splice sites, polypyrimidine tracts, branchpoints, and splicing regulatory elements)^86^. SpliceAI-10k builds on this approach by using SpliceAI scores to systematically predict splicing aberrations (pseudoexonization, partial intron retention, partial exon deletion, exon skipping, and whole intron retention), altered transcript sizes, and consequent amino acid sequences^85^. In order to identify genetic variants to input to SpliceAI-10k, we performed finemapping using SuSiE^87^ for all splicing events identified in the TWAS analysis using the relevant GTEx tissue splicing data with a window of ±100kb around each splicing event. Genetic variants within credible sets were then filtered for those which were within 100:1 log likelihood of also being a CRC risk variant (i.e. genetic variant P-value is within two orders of magnitude from the top genetic variant in the CRC GWAS). For splicing events for which no credible sets were identified, all genetic variants within 100:1 log likelihood of being a CRC risk variant were used. All resulting genetic variants were then input to the SpliceAI-10k calculator.

### Shared causal pathways with known CRC risk factors

To investigate shared causal pathways between our prioritised genes and known CRC risk factors, we performed genetic colocalisation as in our prior analysis. For each of the four previously identified CRC risk factors (BMI, WHR, alcohol consumption, and tobacco use), we performed colocalisation for expression of all genes with robust evidence (i.e. P < Bonferroni threshold in relevant MR analysis and H_4_ > 0.8 in colocalisation analysis) for a causal effect of expression on CRC risk, and the risk factor. We again applied a posterior probability threshold of H_4_ > 0.8 as evidence for a shared causal variant between traits^84^. In such cases, this suggests that there may be a shared causal pathway between expression of the gene and the risk factor. This could be indicative of a mediating role of expression of that gene in the effect of risk factors on CRC risk (e.g. increased BMI may increase expression of the gene which may increase risk of CRC). Alternatively, it may be that expression of the gene influences liability to the risk factor, which then increases risk of CRC through further biological pathways (e.g. if increased expression of the gene increases BMI, which then causes CRC through alternative pathways). We repeated analyses with sex-specific GWAS where data were available as a sensitivity analysis (i.e. for BMI and WHR; see Supplementary table 12 for the sex-specific data sources.

### Statistical analyses

Units of gene expression betas, as outlined by the GTEx consortium, are the result of a normalisation procedure consisting of normalisation between samples using the trimmed mean of M values method^88^, followed by normalisation across samples by inverse normal transformation, and as such the normalised expression units have no direct biological interpretation (see https://gtexportal.org/home/methods for more information)^33^. All analyses were performed using R^89^ (version 4.0.2) or Python^90^ (version 3.9.13, other than the GWAS imputation step of the TWAS analysis which was performed using version 3.5.0). The following R packages were used: for reading in or wrangling data, data.table^91^ (version 1.14.8), dplyr^92^ (version 1.1.1), tidyr^93^ (version 1.3.0), stringr^94^ (version 1.5.0), readxl^95^ (version 1.4.2); for visualisations, ggforestplot^96^ (version 0.1.0), ggrepel^97^ (version 0.9.3), ggplot2^98^ (version 3.4.2), ggpubr^99^ (version 0.6.0), qqman^100^ (version 0.1.8); for colocalisation analyses, coloc^82^ (version 5.1.0.1); for MR analyses TwoSampleMR^101,102^ (version 0.5.5), gwasglue^103^ (version 0.0.0.9000); for compiling LD reference panels, plinkbinr^104^ (version 0.0.0.9000), ieugwasr^105^ (version 0.1.5); for accessing Ensembl databases, biomaRt^106,107^ (version 2.46.3); for finemapping, susieR^87,108^ (version 0.12.35).

## Data and code availability

All data generated by this analysis can be found within the manuscript and supporting information. The CRC GWAS data used in this analysis can be accessed through the GECCO data access process (outlined at https://share.fhcrc.org/sites/gecco/Pages/paper-proposals.aspx).

## Acknowledgements

EH is supported by a Cancer Research UK Population Research Committee Studentship (C18281/A30905), the CRUK Integrative Cancer Epidemiology Programme (C18281/A29019), the Harold Hyam Wingate Foundation, the European Cancer Prevention (ECP) organisation, the European Association for Cancer Research (EACR), and is part of the Medical Research Council Integrative Epidemiology Unit at the University of Bristol which is supported by the Medical Research Council (MC_UU_00032/03) and the University of Bristol. TAO’M is supported by a National Health and Medical Research Council of Australia Investigator Fellowship (Emerging Leadership 2; APP1179170).

The CRC GWAS used in this analysis was provided by: GECCO, which acknowledges funding from the National Cancer Institute, National Institutes of Health, U.S. Department of Health and Human Services (U01 CA137088, R01 CA059045, R01 201407), with genotyping/sequencing services provided by the Center for Inherited Disease Research (CIDR) contract number HHSN268201700006I and HHSN268201200008I, and was funded in part through the NIH/NCI Cancer Center Support Grant P30 CA015704, with scientific Computing Infrastructure at Fred Hutch funded by ORIP grant S10OD028685; the CORECT study which was supported by the National Cancer Institute, National Institutes of Health (NCI/NIH), U.S. Department of Health and Human Services (grant numbers U19 CA148107, R01 CA081488, P30 CA014089, R01 CA197350; P01 CA196569; R01 CA201407; R01 CA242218), National Institutes of Environmental Health Sciences, National Institutes of Health (grant number T32 ES013678), and a generous gift from Daniel and Maryann Fong; and the Colon Cancer Family Registry (CCFR, https://www.coloncfr.org/), which is supported in part by funding from the National Cancer Institute (NCI), National Institutes of Health (NIH) (award U01 CA167551), with support for case ascertainment provided in part from the Surveillance, Epidemiology, and End Results (SEER) Program and the following US state cancer registries: AZ, CO, MN, NC, NH; and by the Victoria Cancer Registry (Australia) and Ontario Cancer Registry (Canada). The CCFR Set-1 (Illumina 1M/1M-Duo) and Set-2 (Illumina Omni1-Quad) scans were supported by NIH awards U01 CA122839 and R01 CA143237 (to GC). The CCFR Set-3 (Affymetrix Axiom CORECT Set array) was supported by NIH award U19 CA148107 and R01 CA81488 (to SBG). The CCFR Set-4 (Illumina OncoArray 600K genetic variant array) was supported by NIH award U19 CA148107 (to SBG) and by the Center for Inherited Disease Research (CIDR), which is funded by the NIH to the Johns Hopkins University, contract number HHSN268201200008I. Additional funding for the OFCCR/ARCTIC was through award GL201-043 from the Ontario Research Fund (to BWZ), award 112746 from the Canadian Institutes of Health Research (to TJH), through a Cancer Risk Evaluation (CaRE) Program grant from the Canadian Cancer Society (to SG), and through generous support from the Ontario Ministry of Research and Innovation. The SFCCR Illumina HumanCytoSNP array was supported in part through NCI/NIH awards U01/U24 CA074794 and R01 CA076366 (to PAN). The content of this manuscript does not necessarily reflect the views or policies of the NCI, NIH or any of the collaborating centres in the Colon Cancer Family Registry (CCFR), nor does mention of trade names, commercial products, or organizations imply endorsement by the US Government, any cancer registry, or the CCFR.

The funders had no role in study design, data collection and analysis, decision to publish, or preparation of the manuscript.

