## Supplementary Note for "Integrating multi-tissue expression and splicing data to prioritise anatomical subsite- and sex-specific colorectal cancer susceptibility genes with therapeutic potential"

**STROBE-MR checklist of recommended items to address in reports of Mendelian randomization studies**^1^ ^2^

| **Item No.** | **Section** | **Checklist item** | **Page No.** | **Relevant text from manuscript** |
| --- | --- | --- | --- | --- |
| 1 | **TITLE and ABSTRACT** | Indicate Mendelian randomization (MR) as the study’s design in the title and/or the abstract if that is a main purpose of the study | 4 | involving Mendelian randomisation |
|  | **INTRODUCTION** |  |  |  |
| 2 | **Background** | Explain the scientific background and rationale for the reported study. What is the exposure? Is a potential causal relationship between exposure and outcome plausible? Justify why MR is a helpful method to address the study question | 5 | no CRC TWAS performed thus far has stratified by anatomical subsite or sex, which are important aspects of CRC development. Additionally, TWAS for CRC have often lacked a causal framework analysis |
| 3 | **Objectives** | State specific objectives clearly, including pre-specified causal hypotheses (if any). State that MR is a method that, under specific assumptions, intends to estimate causal effects | 6, 12 | genes, we performed comprehensive multi-tissue TWAS analyses as outlined in **Figure 1**. We then employed a causal framework performing Mendelian randomization (MR)  Under certain assumptions, MR can provide causal estimates (see **Methods**) |
|  | **METHODS** |  |  |  |
| 4 | **Study design and data sources** | Present key elements of the study design early in the article. Consider including a table listing sources of data for all phases of the study. For each data source contributing to the analysis, describe the following: |  |  |
|  | a) | Setting: Describe the study design and the underlying population, if possible. Describe the setting, locations, and relevant dates, including periods of recruitment, exposure, follow-up, and data collection, when available. | 14 | Summary genetic association data for CRC risk (52,775 cases, 45,940 controls) |
|  | b) | Participants: Give the eligibility criteria, and the sources and methods of selection of participants. Report the sample size, and whether any power or sample size calculations were carried out prior to the main analysis | 14 | (52,775 cases, 45,940 controls) |
|  | c) | Describe measurement, quality control and selection of genetic variants | 16 | We identified genetic instruments as genetic variants which are *cis*-acting (i.e. within 100kb of the gene coding region), strongly associated with gene expression (*P* < 5 × 10^-8^), independent (r^2^ < 0.001), and had an F-statistic > 10 |
|  | d) | For each exposure, outcome, and other relevant variables, describe methods of assessment and diagnostic criteria for diseases | 7 | Briefly, proximal, distal and rectal are mutually exclusive anatomical subsites designated by location of tumour, whereas colon is comprised of proximal colon and distal colon tumours, as well as colon cancer with unspecified location. |
|  | e) | Provide details of ethics committee approval and participant informed consent, if relevant |  |  |
| 5 | **Assumptions** | Explicitly state the three core IV assumptions for the main analysis (relevance, independence and exclusion restriction) as well assumptions for any additional or sensitivity analysis | 15 | The three core assumptions of MR state that: (i) the genetic variant(s) are strongly and robustly associated with the exposure; (ii) there is no confounding of the genetic variant(s)-outcome relationship (e.g., population stratification); (iii) the genetic variant(s) only affect the outcome through their effect on the exposure. |
| 6 | **Statistical methods: main analysis** | Describe statistical methods and statistics used |  |  |
|  | a) | Describe how quantitative variables were handled in the analyses (i.e., scale, units, model) | 18 | Units of gene expression betas, as outlined by the GTEx consortium, are the result of a normalisation procedure consisting of normalisation between samples using the trimmed mean of M values (TMM) method, followed by normalisation across samples by inverse normal transformation, and as such the normalised expression units have no direct biological interpretation (see <https://gtexportal.org/home/methods> for more information) |
|  | b) | Describe how genetic variants were handled in the analyses and, if applicable, how their weights were selected |  |  |
|  | c) | Describe the MR estimator (e.g. two-stage least squares, Wald ratio) and related statistics. Detail the included covariates and, in case of two-sample MR, whether the same covariate set was used for adjustment in the two samples | 16 | Where only a single genetic variant was available, we calculated the Wald ratio to generate effect estimates; where multiple genetic variants were available, an inverse variable weighted (IVW) multiplicative random effects model was used. |
|  | d) | Explain how missing data were addressed |  |  |
|  | e) | If applicable, indicate how multiple testing was addressed | 16 | A Bonferroni-correction was applied to account for multiple testing (*P* < 4.38 × 10^-5^; 0.05/N*G where N is the number of gene-tissue pairs (161) and G is the number of CRC GWAS (overall, male, female, colon, distal, proximal, rectal). |
| 7 | **Assessment of assumptions** | Describe any methods or prior knowledge used to assess the assumptions or justify their validity |  |  |
| 8 | **Sensitivity analyses and additional analyses** | Describe any sensitivity analyses or additional analyses performed (e.g. comparison of effect estimates from different approaches, independent replication, bias analytic techniques, validation of instruments, simulations) |  |  |
| 9 | **Software and pre-registration** |  |  |  |
|  | a) | Name statistical software and package(s), including version and settings used | 18 | “Statistical analyses” section |
|  | b) | State whether the study protocol and details were pre-registered (as well as when and where) |  |  |
|  | **RESULTS** |  |  |  |
| 10 | **Descriptive data** |  |  |  |
|  | a) | Report the numbers of individuals at each stage of included studies and reasons for exclusion. Consider use of a flow diagram |  |  |
|  | b) | Report summary statistics for phenotypic exposure(s), outcome(s), and other relevant variables (e.g. means, SDs, proportions) |  |  |
|  | c) | If the data sources include meta-analyses of previous studies, provide the assessments of heterogeneity across these studies |  |  |
|  | d) | For two-sample MR:  i.  Provide justification of the similarity of the genetic variant-exposure associations between the exposure and outcome samples  ii.  Provide information on the number of individuals who overlap between the exposure and outcome studies | 14 | All almost entirely European ancestry, no individuals likely to overlap. |
| 11 | **Main results** |  |  |  |
|  | a) | Report the associations between genetic variant and exposure, and between genetic variant and outcome, preferably on an interpretable scale |  | Supplementary table 13 |
|  | b) | Report MR estimates of the relationship between exposure and outcome, and the measures of uncertainty from the MR analysis, on an interpretable scale, such as odds ratio or relative risk per SD difference |  | Supplementary tables 4 and 5 |
|  | c) | If relevant, consider translating estimates of relative risk into absolute risk for a meaningful time period |  |  |
|  | d) | Consider plots to visualize results (e.g. forest plot, scatterplot of associations between genetic variants and outcome versus between genetic variants and exposure) |  | Figures 3 and 4 |
| 12 | **Assessment of assumptions** |  |  |  |
|  | a) | Report the assessment of the validity of the assumptions | 7, 12 | Genetic colocalisation analysis assesses whether the same or distinct variant(s) underly the association between two traits many of our MR analyses were restricted to a single SNP, meaning we were unable to employ various “pleiotropy-robust” models to evaluate exclusion restriction assumptions |
|  | b) | Report any additional statistics (e.g., assessments of heterogeneity across genetic variants, such as *I^2^*, Q statistic or E-value) |  |  |
| 13 | **Sensitivity analyses and additional analyses** |  |  |  |
|  | a) | Report any sensitivity analyses to assess the robustness of the main results to violations of the assumptions |  |  |
|  | b) | Report results from other sensitivity analyses or additional analyses |  | Results of TWAS and colocalization throughout results section |
|  | c) | Report any assessment of direction of causal relationship (e.g., bidirectional MR) |  |  |
|  | d) | When relevant, report and compare with estimates from non-MR analyses |  |  |
|  | e) | Consider additional plots to visualize results (e.g., leave-one-out analyses) |  |  |
|  | **DISCUSSION** |  |  |  |
| 14 | **Key results** | Summarize key results with reference to study objectives | 10 | Using this framework, we identified 35 genes with a likely causal association |
| 15 | **Limitations** | Discuss limitations of the study, taking into account the validity of the IV assumptions, other sources of potential bias, and imprecision. Discuss both direction and magnitude of any potential bias and any efforts to address them | 12 | Our analysis aimed to robustly prioritise CRC susceptibility genes specific to sex and anatomical subsite by using multiple tissues alongside a causal framework. However… |
| 16 | **Interpretation** |  |  |  |
|  | a) | Meaning: Give a cautious overall interpretation of results in the context of their limitations and in comparison with other studies | 12 | Our analyses have identified genes with robust evidence for a potential causal role in CRC development, providing insights into its aetiology and presenting tangible opportunities for targeted therapeutic interventions. |
|  | b) | Mechanism: Discuss underlying biological mechanisms that could drive a potential causal relationship between the investigated exposure and the outcome, and whether the gene-environment equivalence assumption is reasonable. Use causal language carefully, clarifying that IV estimates may provide causal effects only under certain assumptions | Throughout discussion | e.g. Cerebral cavernous malformation 2 (CCM2) is a component of the CCM signalling complex, which has a role in regulating several signalling cascades, including progesterone signalling |
|  | c) | Clinical relevance: Discuss whether the results have clinical or public policy relevance, and to what extent they inform effect sizes of possible interventions | 12 | Given the increase in CRC worldwide, understanding the biological mechanisms leading to carcinogenesis is becoming increasingly important. Additionally, as more screening programmes are rolled out globally, opportunities to prevent CRC development in high-risk individuals are also increasing. Therefore, the identification of novel pharmaceutical targets for the prevention and treatment of this disease remains a priority |
| 17 | **Generalizability** | Discuss the generalizability of the study results (a) to other populations, (b) across other exposure periods/timings, and (c) across other levels of exposure | 12 | our analysed were restricted to individuals of predominantly European ancestries, which limits the generalisability of our findings to other populations and contexts |
|  | **OTHER INFORMATION** |  |  |  |
| 18 | **Funding** | Describe sources of funding and the role of funders in the present study and, if applicable, sources of funding for the databases and original study or studies on which the present study is based | 19 | Acknowledgements section |
| 19 | **Data and data sharing** | Provide the data used to perform all analyses or report where and how the data can be accessed, and reference these sources in the article. Provide the statistical code needed to reproduce the results in the article, or report whether the code is publicly accessible and if so, where | 18 | All data generated by this analysis can be found within the manuscript and supporting information, or the online repository on Zenodo |
| 20 | **Conflicts of Interest** | All authors should declare all potential conflicts of interest | 20 | Declaration of interests section |

This checklist is copyrighted by the Equator Network under the Creative Commons Attribution 3.0 Unported (CC BY 3.0) license.

1.	Skrivankova VW, Richmond RC, Woolf BAR, Yarmolinsky J, Davies NM, Swanson SA, et al. Strengthening the Reporting of Observational Studies in Epidemiology using Mendelian Randomization (STROBE-MR) Statement. JAMA. 2021;under review.

2.	Skrivankova VW, Richmond RC, Woolf BAR, Davies NM, Swanson SA, VanderWeele TJ, et al. Strengthening the Reporting of Observational Studies in Epidemiology using Mendelian Randomisation (STROBE-MR): Explanation and Elaboration. BMJ. 2021;375:n2233.
