## Supplementary figures and images for "Integrating multi-tissue expression and splicing data to prioritise anatomical subsite- and sex-specific colorectal cancer susceptibility genes with therapeutic potential"

### Supplementary Figure 1

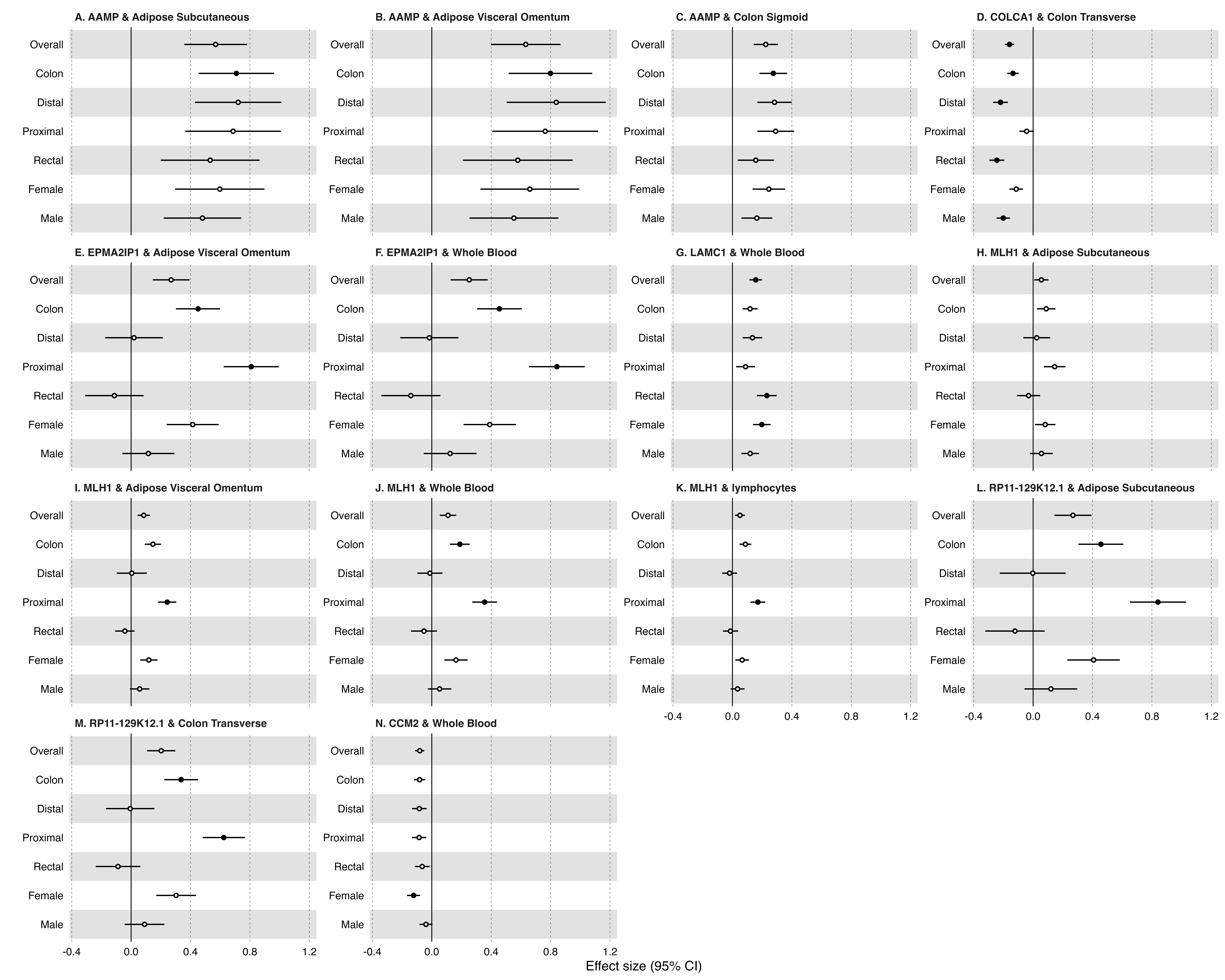

### Supplementary Figure 2

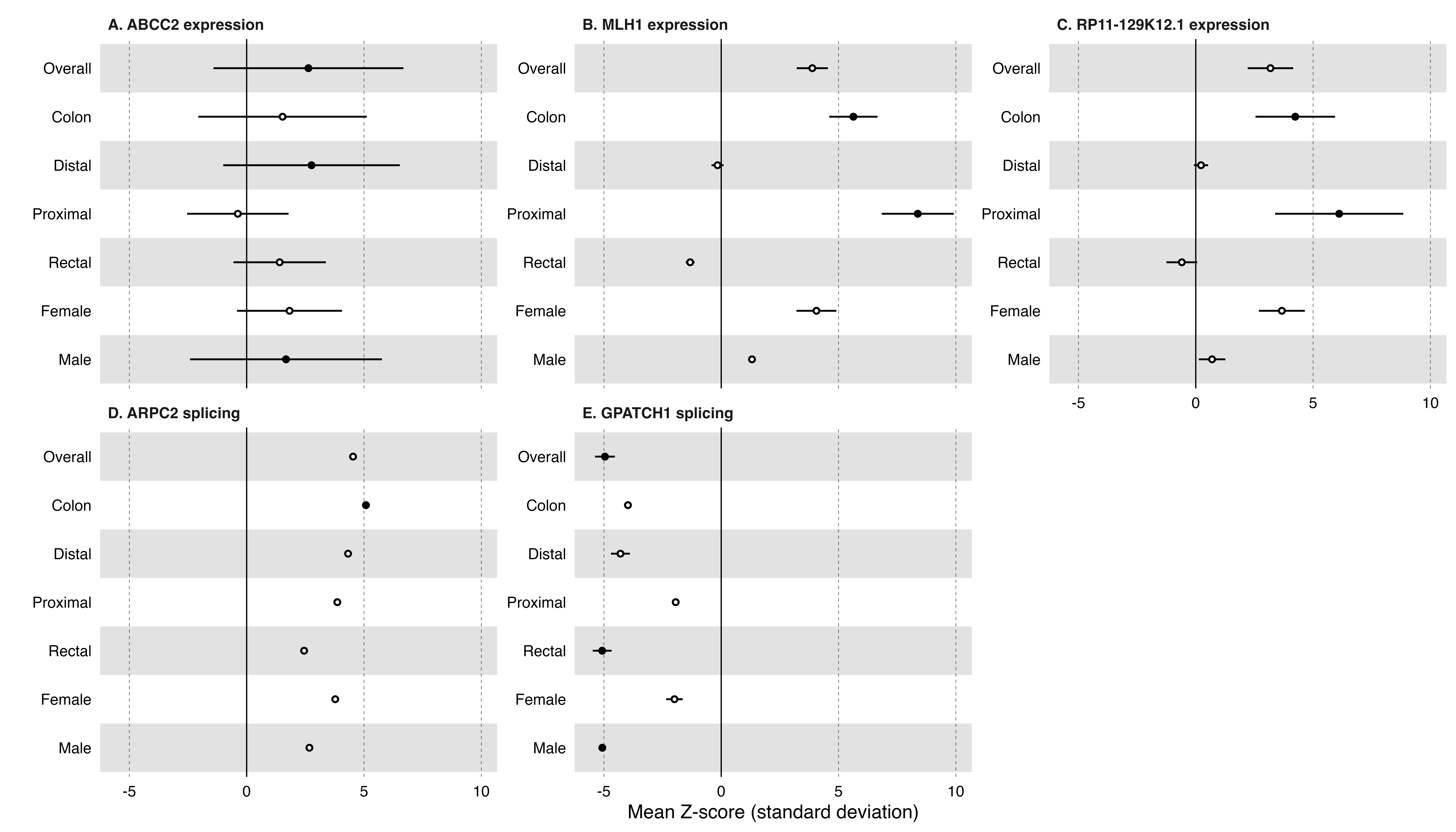
